# Dosimetry-Driven Alpha Emitter Selection for Radioligand Therapy: Is daughter translocation a significant safety concern?

**DOI:** 10.1101/2025.08.08.25333110

**Authors:** Benjamin Fongenie, Nathaniel Scott, Nicolas Chouin, Ana M. Denis-Bacelar, Daniel J Stevens

## Abstract

**PURPOSE:** The first long-term safety data with a ^212^Pb-labelled radiopharmaceutical reported significant renal adverse events after more than two years of follow-up. Understanding the pharmacokinetic drivers of dose delivery for alpha emitting radioligand therapy (RLT) will be critical to optimizing molecules. We aimed to model and compare the dosimetry and therapeutic index (TI) of a single pharmaceutical, rhPSMA-10.1, when labeled with either ^225^Ac or ^212^Pb, using human pharmacokinetic data.

**METHODS:** Dosimetry data from a Phase 1 trial of ^177^Lu-rhPSMA-10.1 in 13 mCRPC patients was used to generate time–activity curves for tumors and select organs-at-risk. These curves were used to model the dosimetry of ^225^Ac-rhPSMA-10.1 and ^212^Pb-rhPSMA-10.1 by substituting the physical half-life and decay properties of ^177^Lu with those of the alpha-emitters. Absorbed doses and TIs were calculated. The potential impact of daughter radionuclide translocation on organ doses and TIs was also modeled.

**RESULTS:** To deliver 5Gy_(RBE5)_ absorbed dose to tumors, a ∼29-fold higher administered activity of ^212^Pb was required compared to ^225^Ac (131 MBq versus 4.6 MBq, respectively). At this tumor dose, the activity of ^212^Pb resulted in 2.5-fold and 2.2-fold higher absorbed doses to the kidneys and salivary glands, respectively, compared with ^225^Ac. Consequently, ^225^Ac demonstrated a substantially improved TI, with a ∼3-fold higher tumor:kidney dose ratio (9.85 versus 3.36) and a ∼2.2-fold higher tumor:salivary gland ratio (15.9 versus. 7.1). Even when modeling a worst-case scenario for daughter translocation, ^225^Ac maintained a superior TI. Importantly, based on the pharmacokinetics of this drug, to achieve 120Gy_(RBE5)_ absorbed dose to tumors would require the delivery of 12.1Gy_(RBE5)_ and 35.7Gy_(RBE5)_ to the kidneys for ^225^Ac and ^212^Pb respectively. Modeling daughter translocation, these values become 39.1Gy_(RBE5)_ and 114.3Gy_(RBE5)_ respectively which may help to explain recently reported safety data from ^212^Pb-labelled RLT.

**CONCLUSIONS:** The physical half-life of ^225^Ac is better suited to the pharmacokinetic profile of rhPSMA-10.1 than the shorter half-life of ^212^Pb. This results in substantially enhanced TI for ^225^Ac-rhPSMA-10.1, permitting the delivery of markedly lower absorbed doses to organs-at-risk for a fixed tumor dose. Importantly, labelling with ^212^Pb may lead to very high renal absorbed radiation doses driven predominantly by demetallation.

**GRAPHICAL ABSTRACT:** 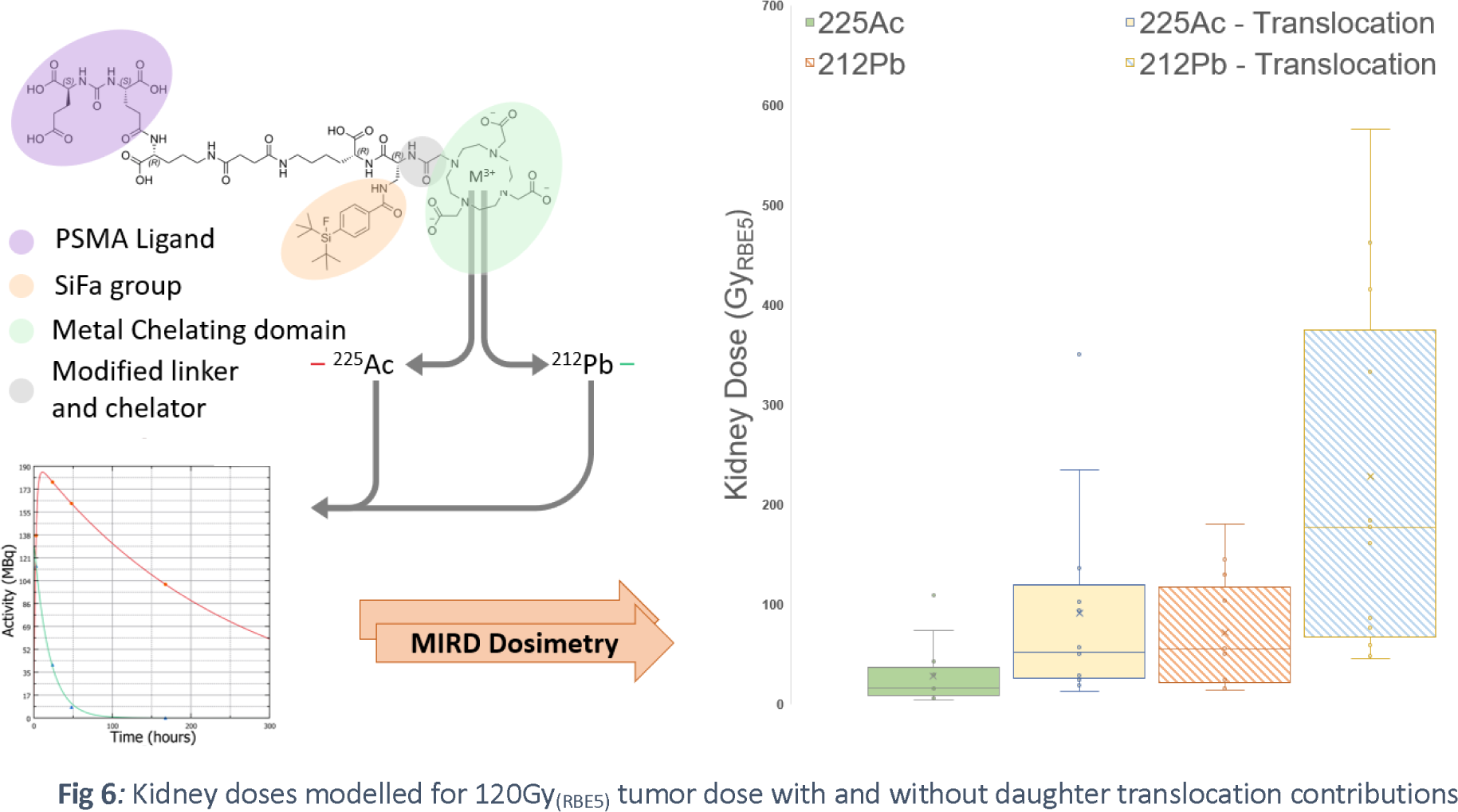

## INTRODUCTION

Imaging emissions secondary to radioligand therapy (RLT) offers a unique ability to provide quantitative estimates of the exposure to radioactivity of each tissue in humans, permitting the calculation of estimated absorbed doses to tumors and normal tissues. Currently, a range of novel RLTs are being explored for numerous targets using both beta and alpha-emitters. Specifically, there is significant interest in alpha-emitting radionuclides due to their high linear energy transfer and short range, which causes dense complex ionization tracks when damaging DNA [1]. However, performing quantitative radiation dosimetry with the two most available alpha emitters (^225^Ac and ^212^Pb) is challenging given their emission characteristics and complex decay chains (Figure 1). In addition, the dislocation of daughter radionuclides for both alpha-emitters and subsequent deposition around the body - “translocation” - complicates the estimation of absorbed dose.

**Fig 1:**
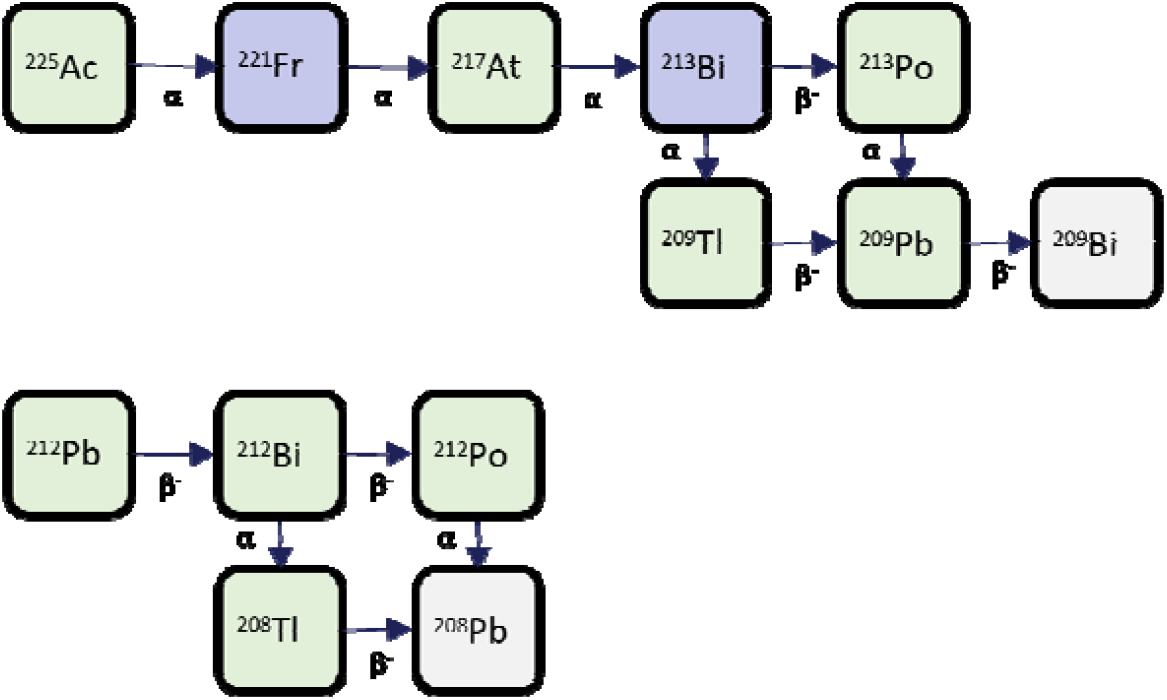
Decay chains for 225Ac and 212Pb

The use of a “surrogate” radionuclide with excellent imaging properties (e.g. ^177^Lu, ^111^In) is a common strategy to assess the pharmacokinetics and radiation dosimetry of a novel RLT prior to committing to larger trials exploring safety and efficacy. The data generated can also be used to assess which alpha emitting radionuclides may offer the best balance of dose delivered to tumors and to normal tissue, despite the inability to make direct observations with the radionuclides under consideration due to lack of suitable emissions for imaging.

Fundamentally, it is accepted that an ideal agent would deliver all of its absorbed dose to the tumor and none to normal tissue. In reality, it is unavoidable that some radiation dose will be delivered to normal tissue via the expression of the target of interest (“on-target”) and then through exposure to radioactivity from the distribution, metabolism and excretion that comes with delivering a RLT parenterally (“off-target”). Given this, we can define a putative Therapeutic Index (TI) as the ratio of absorbed dose to the tumor versus absorbed dose to key organs-at-risk. Currently, there is significant scientific debate regarding which radionuclides are optimal for therapeutic use [2]. These discussions often center around the physical properties of the radionuclide in relation to radiobiology, considering biologically effective dose, crossfire effects and range in tissue, as well as the physical half-life [3].

However, the interplay between physical radionuclide half-life and the specific pharmacokinetics of a novel molecule has, surprisingly, rarely been modelled or documented in terms of eventual optimization of therapeutic potential [4]. Here, using a novel human dataset from a recently performed Phase 1 trial with ^177^Lu-rhPSMA-10.1 (NCT05413850) we explore the use of two commercially available alpha-emitting radionuclides (^225^Ac and ^212^Pb) and the impact which switching out the ^177^Lu for either of them would have on the estimated absorbed doses and the TI [5, 6].

## METHODS

### Dosimetry

Using dosimetry data from a Phase 1 trial of novel RLT, ^177^Lu-rhPSMA-10.1, in 13 patients with metastatic castration-resistant prostate cancer [NCT05413850], time–activity curves were generated for the first treatment cycle (Cycle 1) for each of the 13 participants [7]. Patient details are described in Table 1. These data were acquired with 4 time-point SPECT/CT scans at 3 hours, 24 hours, 48 hours and 168 hours post-administration. In addition, radioactive blood samples were acquired at 0.5-, 1.5-, 4-, 24- and 48-hours post-administration. The scanners & gamma counters used to acquire these data were calibrated prior to imaging. From the SPECT/CT scans, activities were measured within volumes of interest in tumors and normal tissues at each of these time-points and cumulative activities calculated for tumors and organs-at-risk, similar to Nagarajah *et al* [7].

**Table 1.** Patient and lesion characteristics as previously reported *[7]*.

| Patient | Number of lesions | Lesion types | Average lesion mass (g) | Mass range (g) |
| --- | --- | --- | --- | --- |
| 1 | 5 | Bone | 90.69 | 3.4–427.5 |
| 2 | 2 | Bone | 17.01 | 13.7–20.3 |
| 3 | 4 | Mixed | 10.29 | 2.9–20.7 |
| 4 | 5 | Mixed | 12.62 | 7.5–17.6 |
| 5 | 1 | Bone | 60.30 | N/A |
| 6 | 5 | Bone | 25.21 | 3.2–107.0 |
| 7 | 4 | Lymph node | 6.06 | 2.2–14.8 |
| 8 | 5 | Bone | 30.18 | 8.0–68.8 |
| 9 | 5 | Bone | 32.96 | 18.6–48.2 |
| 10 | 2 | Lymph node | 52.98 | 6.5–99.5 |
| 11 | 5 | Mixed | 13.31 | 6.2–22.1 |
| 12 | 5 | Mixed | 16.10 | 9.6–26.1 |
| 13 | 5 | Mixed | 4.77 | 2.8–9.1 |

**Table 2.** Assumptions for modelling. -*Tumor S-values shown for a ssumed 10 g tumor [8].

| Organ | Fit function | <sup>225</sup>Ac S-value | <sup>212</sup>Pb S-value |
| --- | --- | --- | --- |
| Tumor* | $A \cdot \exp(-(\lambda_1 + \lambda_{phys})) - A \cdot \exp(-(\lambda_2 + \lambda_{phys}))$ | 1608 mGy/MBqh | 679 mGy/MBqh |
| Kidney | $A \cdot \exp(-(\lambda_1 + \lambda_{phys}))$ | 38.9 mGy/MBqh | 3.91 mGy/MBqh |
| Salivary gland | $A \cdot \exp(-(\lambda_1 + \lambda_{phys})) - A \cdot \exp(-(\lambda_2 + \lambda_{phys}))$ | 184 mGy/MBqh | 18.2 mGy/MBqh |

For the purposes of modelling, dosimetry data acquired above was corrected for physical decay of ^177^Lu before applying the physical decay constant for ^225^Ac and ^212^Pb. Thus, the activity within the regions of interest of delineable normal tissues and tumors from all imaging time points as well as activities measured in blood samples were extrapolated from the 6.7-day half-life of ^177^Lu to the half-life of 9.9-days of ^225^Ac and 10.64 hours of ^212^Pb. In this way, effective half-lives for tissues were obtained assuming identical biological half-lives between different radionuclides, given the pharmaceutical was unchanged. Cumulative activities were calculated by integrating under these curves, and absorbed doses were calculated according to S-values calculated using the MIRD formulation (MIRDcalc internal dose calculation software v20220504) [8]. Therapeutic indices were then determined as the ratios of absorbed doses (Gy/Gy) as shown in Equation 1:

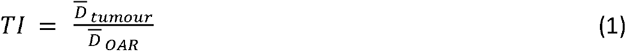

Where 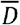 is the absorbed dose to the *tumor* or *organ-at-risk (OAR)*.

As this was a novel prostate-specific membrane antigen (PSMA)-targeted RLT, organs-at-risk were selected as those which are of significant concern due to known dose deposition from the expression of PSMA and elimination routes. These included the salivary glands and the kidneys. The data were plotted using LabPlot Version 2.10.0, a C++ compiled data plotting software. Data was fitted with mono- or bi-exponential functions selected using statistical tests as recommended by Ivashchenko et al [9].

The statistical results from model fit testing can be found in the Supplemental Table 1. These fits were then integrated to a 720-hour cut-off using LabPlot2’s integral analysis tool (Simpson’s 3-point method). For calculation of absorbed doses, the S-values listed in Table 1 were used for the associated tissues and the commonly used relative biological effectiveness value of 5 (RBE5) was used to account for the increased cytotoxicity of alpha particles versus gamma or beta radiation.

### Translocation

For radiopharmaceuticals labelled with ^225^Ac or ^212^Pb it is understood that decays of the parent radionuclide can lead to dislocation of daughters and subsequent untargeted deposition of “free” daughters throughout the body, colloquially known as “translocation”. Importantly, normal organ doses and therefore therapeutic ratios are sensitive to dislocation and translocation of daughters. In the case of ^225^Ac, dislocation of daughters happens in ∼100% of decays due to the recoil effect, whereas in the case of ^212^Pb dislocation occurs via chemical interactions due to changes in charge, often referred to as demetallation. This demetallation is thought to occur anywhere from ∼36% to ∼5% of the time depending on the study and source [5, 10, 11]. Data from human trials has confirmed that ^212^Pb demetallation occurs in PSMA-targeting radioligands [12]. The impact of dislocation and translocation of daughters on absorbed doses is difficult to determine, but data presented using a different novel ^225^Ac-labelled PSMA targeting small molecule imply almost no translocation occurs from or to target lesions in mice [13, 14]. In this analysis by Zitzmann-Kolbe *et al*, biodistribution data was collected from tumors, kidneys and salivary glands and a range of timepoints. The samples were measured in a gamma counter immediately following sacrifice and many hours later, when secular equilibrium with daughters was in place. As such the excess or missing ^221^Fr and ^213^Bi versus expected equilibrium activities could be measured. The lack of translocation to or from the tumor suggests the majority of translocation happens due to decays occurring in plasma. These data resulted in increase to the kidney dose by a factor of ∼3.2 and salivary dose by a factor of 4, with no change in tumour dose. Here-in these factors are referred to as “dose-shift”. While it is possible to simulate biodistribution and dosimetric impact from daughters using ICRP biokinetic models, previous studies have demonstrated that these models are highly sensitive to compartment assumptions [15]. Therefore, as an alternative approach, here the dose-shift measured in mice is used as a base case.

Using the assumption this absorbed dose increase is a result of decays occurring in free plasma, and it is accurate for ^225^Ac, the known plasma half-life of ^177^Lu-rhPSMA-10.1 acquired with radioactive blood sampling on phase 1 patients was used to calculate the proportion of total potential decays which occur in plasma for both ^225^Ac-rhPSMA-10.1 and ^212^Pb-rhPSMA-10.1. This was done by first adjusting for the physical half-lives of the alpha-emitters to simulate plasma-washout curves. Integrating under these curves out to 7 days gives the cumulative activity in plasma 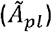 which gives the total number of decays occurring in plasma, while integrating under physical decay gave the total potential decays injected 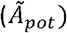:

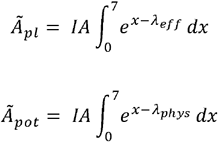

Where *IA* is the simulated injected activity, *x* is the time in days and *λ*_*eff*_ is the effective decay constant of the radioligand in plasma and *λ*_*phys*_ is the physical decay constant of the radionuclide in question. Seven days was selected to coincide with contamination restrictions on ^177^Lu-rhPSMA-10.1 and therefore expected to capture the entirety of washout period. We can then define further variables:

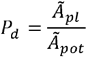

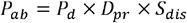

Where *P*_*d*_ is the proportion of potential injected decays occurring in plasma, *P*_*ab*_ is the proportion of potential injected absorbed dose dislocated in plasma, *D*_*pr*_ is the proportional dose contribution from progeny (approximately 0.75 for ^225^Ac and 0.98 for ^212^Pb) and *S*_dis_ is the proportion of decays that lead to dislocation (1 for ^225^Ac, assumed demetallation rate for ^212^Pb).

If we then assume that the dose-shift detected by Zitzmann-Kolbe et al is proportional to *P*_*ab*_ we can identify the point at which *P*_*ab*_ is identical for both ^212^Pb and ^225^Ac as the point at which the dose-shift would be equal. We can also use the relative differences of *P*_*ab*_ to scale the dose-shift of ^212^Pb accordingly for various assumed demetallation rates. For example, if *P*_*ab*_ of ^212^Pb is 2x that of ^225^Ac the dose-shift will be proportional to 2x daughter dose contribution, i.e. for kidney 2 x (2.2 : 1) = 4.4 : 1 ratio gives a dose shift factor of 5.4. This also relies upon the assumption that the uptake into various organs of free ^213^Bi and free ^212^Bi is identical. It should be noted that as the half-life of ^212^Bi is longer than that of ^213^Bi (60.5 vs 45.6 mins) this will likely underestimate the amount of ^212^Bi decaying in key normal tissues like kidney and salivary glands. No adjustments accounting for potential injected free ^221^Fr/^213^Bi were made, given that the majority of ^213^Bi in formulation will be chelated by DTPA and rapidly excreted with minimal dose delivery, and that the contribution from free ^221^Fr is likely negligible.

## RESULTS

An example of fits for tumor and kidney can be found in Figure 2. The results demonstrate that in order to deliver a 5Gy_(RBE5)_ absorbed dose to tumors, a considerably higher (approximately 29-fold) administered activity of ^212^Pb is required than of ^225^Ac (Figure 3). The mean (± standard deviation) results being 131 (± 106) MBq (range: 27–293 MBq) and 4.6 (± 4.4) MBq (range: 0.7–11.3 MBq), respectively. This approximately 29-fold difference is the result of the notably shorter physical half-life leading to a short effective half-life in tumor, which reduces the area under curve and therefore time-integrated cumulative activity for equivalent administered activities. In addition, compared to ^225^Ac, the S-value is notably smaller due to fewer alpha emissions per decay for ^212^Pb.

**Fig 2.**
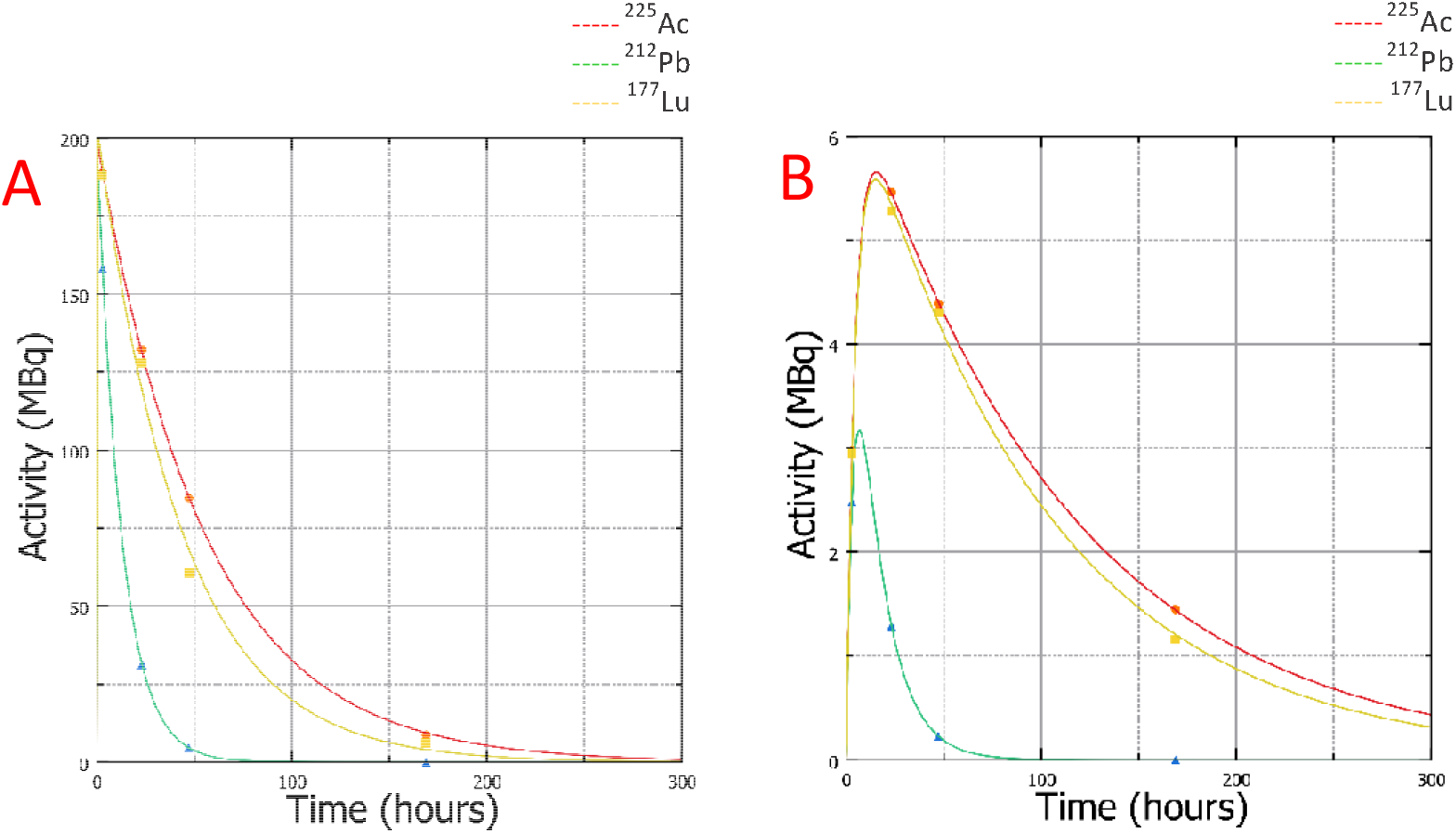
Example tumor and kidney time–activity curves with best-fits. Panel A shows kidney TACs, Panel B shows tumor TACs

**Fig 3:**
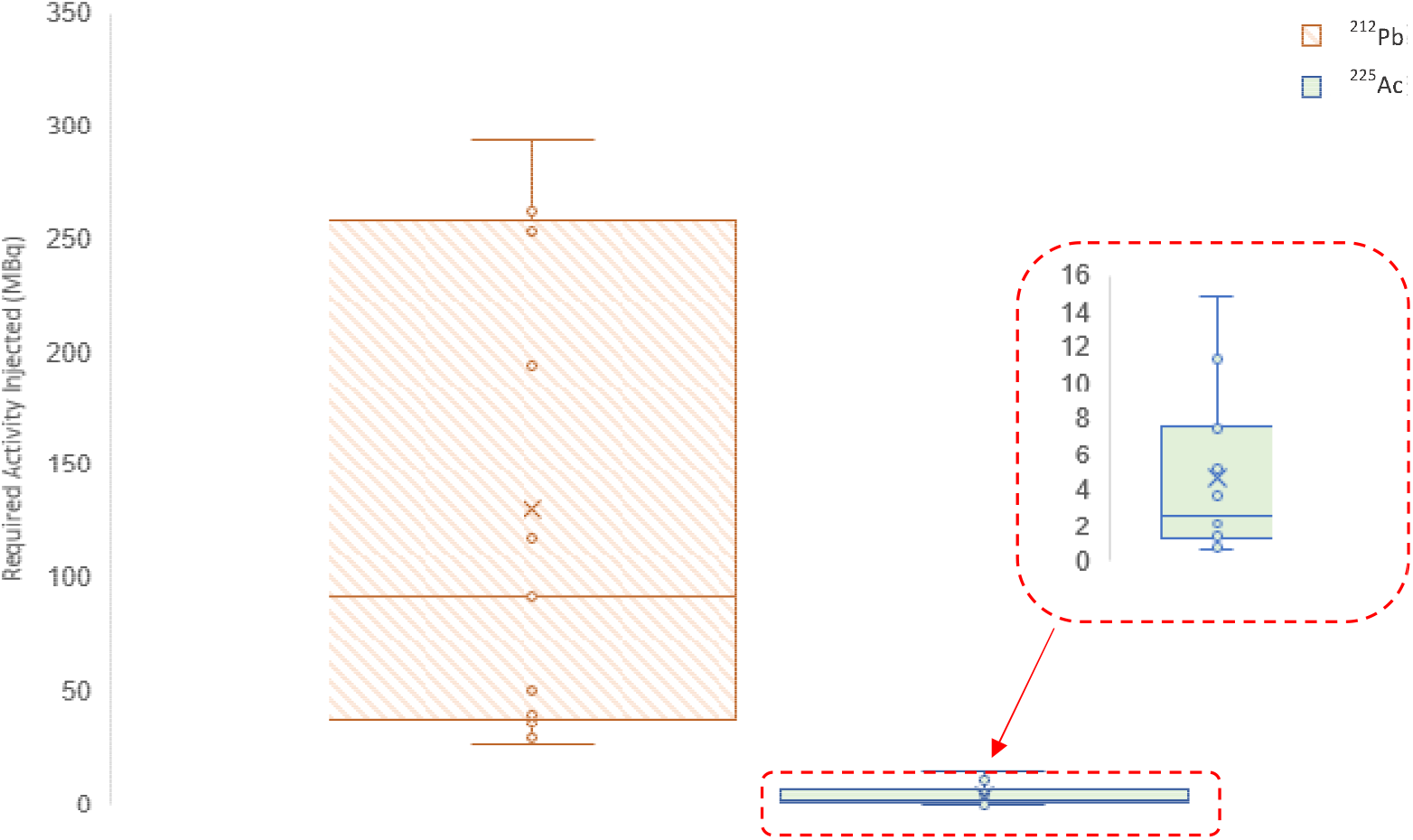
Required activity injected in MBq to achieve an average absorbed tumor dose of 5Gy(RBE5).

Assuming no translocation and a target of 5Gy_(RBE5)_ delivered to the tumor, the absorbed doses delivered to the kidney and salivary glands are substantially different between radionuclides (Figure 4). The higher administered activity of ^212^Pb required for the 5Gy_(RBE5)_ to the tumor results in a 2.5-fold and 2.2-fold higher delivery of dose to the kidney and salivary glands, respectively. The absorbed dose delivered to tumors for a 35Gy_(RBE5)_ dose to kidneys is 2.9-fold higher for ^225^Ac than ^212^Pb (Figure 3c). These normal-organ absorbed doses also lead to significantly improved TIs for the ^225^Ac-labelled radiopharmaceutical versus the ^212^Pb-labelled one. The mean (± standard deviation) tumour:kidney absorbed dose ratios were 9.85 (± 8.49) and 3.36 (± 2.79), respectively, and for the tumour:salivary absorbed dose ratio they were 15.9 (± 12.5) and 7.06 (± 5.76), respectively.

**Fig 4.**
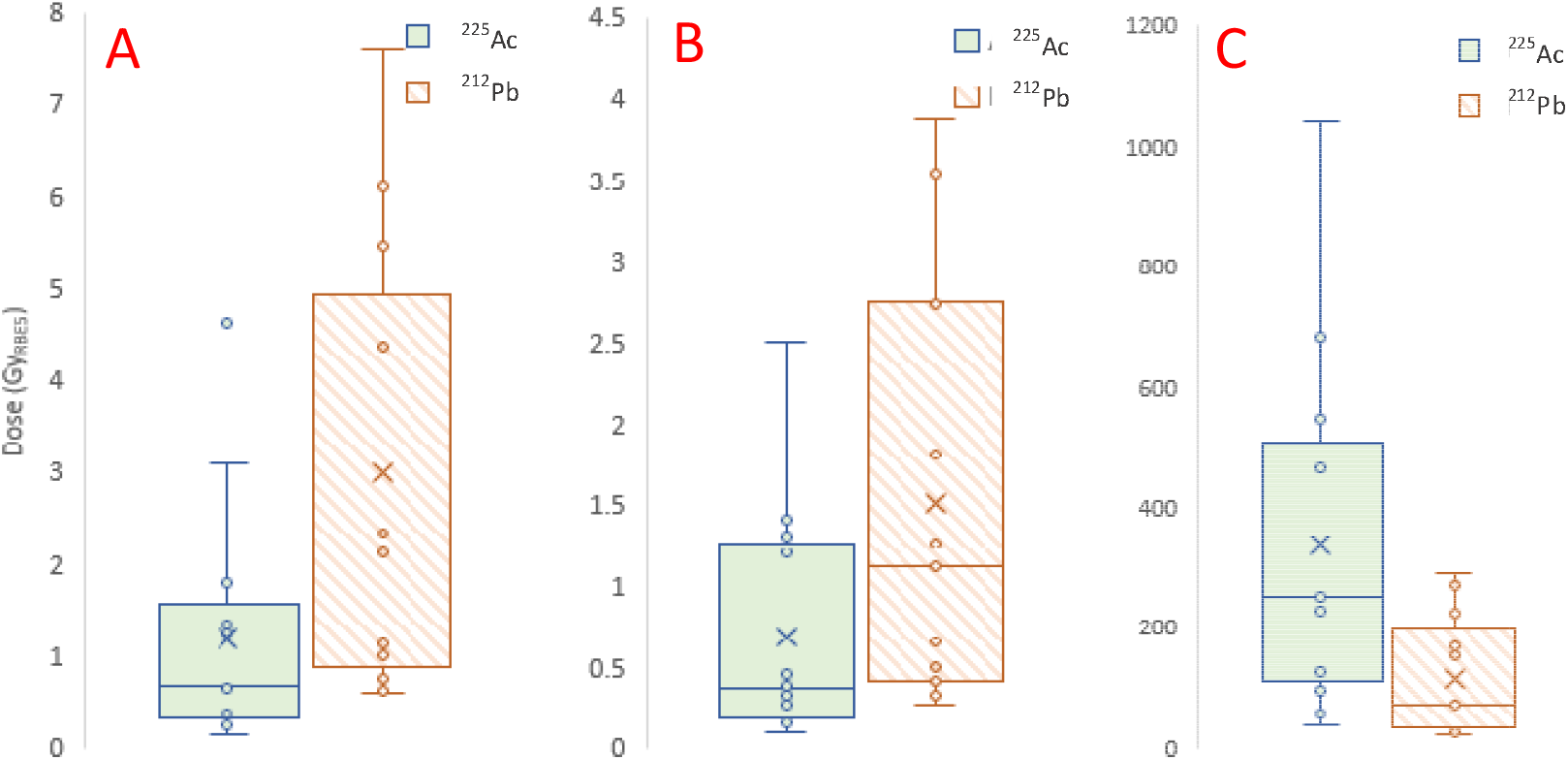
Modelled absorbed doses. Panel A shows the representative kidney dose for 5Gy_(RBE5)_ tumor dose, panel B shows the salivary gland dose for a 5Gy_(RBE5)_ tumor dose, and panel C shows the achievable tumor dose for a 35Gy_(RBE5)_ kidney dose.

Due to the faster decay rate for ^212^Pb and the greater proportion of absorbed dose delivered by ^212^Pb daughters (approximately 98% versus 75% for ^225^Ac) if 11% of ^212^Pb decays lead to demetallation, this results in the same *P*_*ab*_ for ^212^Pb-rhPSMA-10.1 as for ^225^Ac-rhPSMA-10.1, therefore an equal level of dose increase as occurs with ^225^Ac. Given the range and uncertainty of demetallation rates for ^212^Pb complexes, we used a value of 11% for simplicity in the example translocation doses given in the relevant figure (Figure 5). However, we also investigated the impact of translocation for a “worst case” demetallation rate of 36%, as was demonstrated for DOTA chelators by Mirzadeh *et al*, given similar DOTA family chelators are in some cases being investigated for ^212^Pb radioligands in clinical trials [11]. Given the assumed dose-shift for both radionuclides is identical in the translocation modelling as determined by the selected demetallation rate of ^212^Pb being 11%, the difference between TI for each remains the same. The mean (± standard deviation) tumor:kidney absorbed dose ratios in this scenario are 3.08 (± 2.65) and 1.05 (± 0.87) for ^225^Ac and ^212^Pb, respectively. In order for ^212^Pb to match the ^225^Ac TI under these assumptions when accounting for translocation, less than 1% of ^212^Pb decays must lead to demetallation events. Conversely, with 0% ^212^Pb demetallation, the tumor:kidney ratios of ^225^Ac-rhPSMA-10.1 would be better than ^212^Pb-rhPSMA-10.1 if the kidney dose increase due to ^225^Ac daughter translocation was a factor of 2.93 or less.

**Fig 5.**
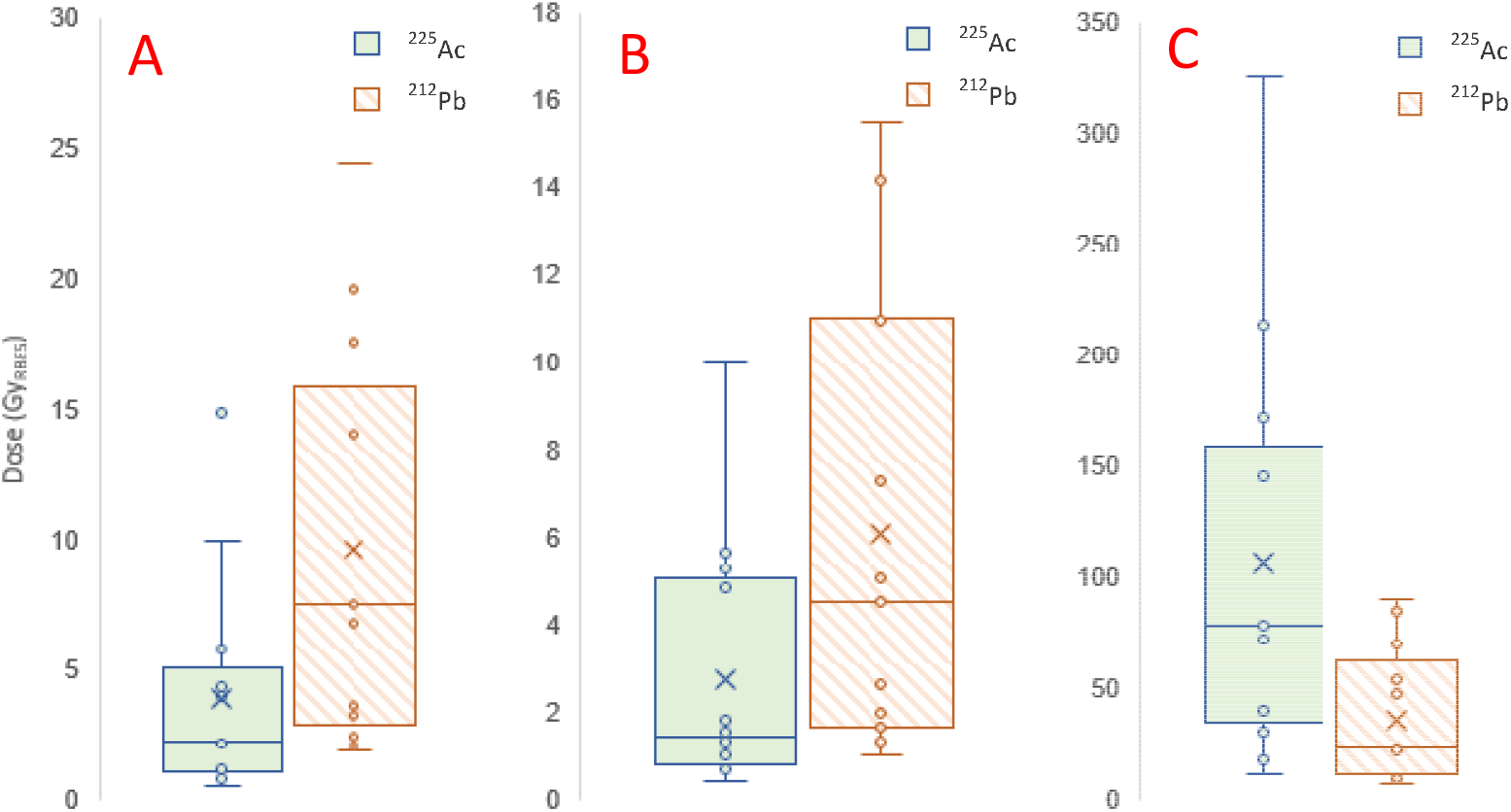
Modelled translocation doses. Panel A shows the kidney dose for 5Gy_(RBE5)_ tumor dose with translocation, panel B shows the salivary dose for 5Gy_(RBE5)_ tumor dose with translocation, and panel C shows the tumor dose for a 35Gy_(RBE5)_ kidney dose.

In the event that 36% of ^212^Pb decays lead to ^212^Bi dislocation, the tumor:kidney absorbed dose ratio for ^212^Pb-rhPSMA-10.1 would be lowered to 0.39. The comparison is shown in supplemental Figure 1.

A systematic review of the only approved PSMA targeted RLT (Pluvicto/^177^Lu-vipivotde tetraxetan) reports an average absorbed tumor dose of 4.04Gy/GBq [16]. Assuming an average of 4 out of a maximum of 6 cycles allowed within the product license, we can assume an average cumulative radiation dose to prostate cancer lesions of ∼120Gy. The impact of attempting to deliver 120 Gy_(RBE5)_ to tumors can be seen in the graphical abstract, leading to kidney doses of 28.67 (±31.04) and 71.23 (±53.43) Gy_(RBE5)_ for ^225^Ac and ^212^Pb respectively without translocation and 91.73 (±99.35) and 227.92 (±170.99) Gy_(RBE5)_ with the translocation assumptions used in this modeling.

The calculated absorbed dose coefficients for kidney and lesions without translocation for ^225^Ac-rhPSMA-10.1 compare favorably with those calculated by Liubchenko *et al* for ^225^Ac-PSMA-I&T: 0.27 (±0.08) Gy_RBE5_ /MBq vs 0.17 (±0.06) Sv_RBE5_/MBq for kidney, 2.6 (±2.45) Gy_RBE5_/MBq vs 0.36 (±0.1) Sv_RBE5_/MBq for lesions (rhPSMA-10.1 vs I&T) [17]. Similarly comparing ^212^Pb-rhPSMA-10.1 to ^212^Pb-CA012 absorbed doses calculated by Kratochwil *et al*: 0.0243 (±0.007) Gy_RBE5_/MBq vs 0.049 Sv_RBE5_/MBq for kidney, 0.08 (±0.06) Gy_RBE5_/MBq vs 0.132 Sv_RBE5_/MBq for lesions (rhPSMA-10.1 vs CA012) [18].

## DISCUSSION

Using the measured pharmacokinetics of ^177^Lu-rhPSMA-10.1 in humans, the impact of labelling the pharmaceutical with ^225^Ac versus ^212^Pb was modelled. Due to the short physical half-life and decay properties of ^212^Pb, notably lower activities of ^225^Ac are required compared with ^212^Pb to achieve clinically meaningful tumor doses. In order to deliver an equivalent dose to tumor, approximately 29-fold more radioactivity is required when considering ^212^Pb compared with ^225^Ac. Even if translocation of daughters is ignored, substantially improved TIs are achieved using ^225^Ac versus ^212^Pb (3-fold higher) due to the long retention in tumor and rapid washout from normal organs of the radiopharmaceutical favoring the longer half-life of ^225^Ac. It should be noted that the decrease in tumor dose for identical kidney doses switching from ^225^Ac to ^212^Pb is not equal to the increased kidney dose for equivalent tumor dose which is a mathematical feature resulting from the method of averaging. When translocation is modelled, unless it is possible to achieve a demetallation rate of <1% of ^212^Pb decays, ^225^Ac continues to offer an improved TI.

The impact of translocation for both radionuclides is based on data derived from a radiopharmaceutical with significantly longer plasma half-life than rhPSMA-10.1 [13]. Therefore, normal organ doses from decays occurring in plasma are likely overestimated and the values presented here can be considered a “worst case scenario” for alpha-labelled rhPSMA-10.1. On the other hand, the contribution of translocation from daughters for antibodies, which have significantly longer plasma half-lives than small molecules, such as J591, is likely to be far greater than the contributions estimated here.

The results of this modelling demonstrate the intrinsic importance of combining pharmacokinetics and radiation dosimetry with radionuclide selection. Ideally, these results would be available prior to the selection of the alpha-emitting radionuclide which might be taken into human clinical trials. Importantly, recently reported clinical data from a ^212^Pb labelled molecule targeting the somatostatin receptor suggested a significant signal for renal toxicity with more than 2 years of follow up data. More than 1 in 3 patients experience some sort of adverse renal event (37%) with 17% experiencing G3 or above adverse renal events [19]. Whilst it is unclear to what extent this is a target specific phenomenon, this safety signal correlates with the findings of this modelling work and supports the idea that demetallation may be driving the deposition of significant renal absorbed radiation dose.

There are some potential limitations to this work. Integrating TACs to 720 hours may introduce bias against ^225^Ac given a small proportion of dose may be delivered after this time-point in exceptional circumstances given its long half-life. It should also be noted that for absorbed dose estimates use an RBE factor for alpha radiation versus gamma or beta radiation. The RBE for alpha used in calculations is 5 and is based on historical in vitro observations [20]. However, there are wide ranges reported for this value and the biological applicability remains unknown in humans [21]. Although, even if this RBE value were not applied, the estimates for activity administered for both ^225^Ac and ^212^Pb would likely increase similarly for both radionuclides and the TIs would remain unchanged. Another aspect of radiobiology not accounted for in this modelling is the impact of dose-rate on biologically effective dose. Dose-rate impact from alpha radiation on the biologically effective dose is currently poorly understood but the literature broadly supports the concept that given the clustered complex ionization tracks created by alpha particles, increasing the dose rate is unlikely to meaningfully increase the proportion of cells killed. The literature proposes that the “single-hit” nature of alpha particle damage means that the ultimate fate of the cell is determined by the total dose received, not the timing of its delivery [22, 23, 24, 25]. However, the majority of these data are derived from non-physiologically relevant in vitro studies and further work in radiobiology is required to understand what, if any, impact dose rate has in human patients with metastatic cancer. It is worth noting that if higher dose rates with alpha particles do lead to a greater proportion of cells killed, it may create a higher risk of toxicity in key normal organs given that those dose rates must also apply to binding target receptor expressed in healthy tissue which is highest in the early hours after injection.

Another limitation of this work is the presumed identical pharmacokinetics of an alpha-emitter labelled version of rhPSMA-10.1, except for physical half-life. It is known that switching ^177^Lu for ^225^Ac causes small chemical changes in the molecule so it may be possible that this would impact the pharmacokinetics beyond the physical half-life difference. However, pre-clinical biodistribution data imply similar behavior between the two and it is reasonable to suppose this may extend to a ^212^Pb-labelled version for the purposes of modelling radiation dosimetry [26].

Additionally, the assumption that dislocated ^213^Bi and ^212^Bi behave identically may be overly simplistic. The dose-shift results relied upon in this paper are generated in mice and so these may not be applicable to humans. The assumption that the proportion of potential absorbed dose dislocated in plasma (*P*_*ab*_) is proportional to dose-shift may also be overly simplistic, for example if there is a significant saturation effect in normal organs. Although, it is difficult to hypothesize that the uptake of free ^212^Bi and ^213^Bi in the kidney could become “saturated” given their status as single atoms. Finally, while we have attempted to fairly model for translocation, we have not accounted for the presence of free ^221^Fr upon administration of the ^225^Ac formulation.

Although preliminary calculations imply an extremely small contribution to normal tissue dose from this phenomenon, including these contributions would add to the accuracy of the modeling [15].

## Conclusion

Based on modeling using human pharmacokinetic data from ^177^Lu-rhPSMA-10.1, we have demonstrated that in this radiopharmaceutical the choice between ^225^Ac and ^212^Pb as alpha-emitters has a profound impact on the agent’s TI. The shorter physical half-life and decay properties of ^212^Pb are poorly matched to the observed pharmacokinetics, leading to a requirement for ∼29-fold higher administered activity to achieve the same tumor dose as ^225^Ac. This mismatch results in a significantly poorer TI for ^212^Pb, with off-target doses to the kidneys and salivary glands approximately 2-to 3-fold higher than with ^225^Ac. Even when daughter radionuclide translocation is considered, ^225^Ac maintains its dosimetric advantage unless the demetallation rate of ^212^Pb is less than 1%, a level not currently supported by any published clinical literature. Our results suggest that for a molecule with a similar pharmacokinetic profile to rhPSMA-10.1, ^225^Ac is the preferred alpha-emitter, offering a more favorable balance of tumor-to-normal tissue dose. This study underscores the fundamental principle that radionuclide selection should not be made in isolation but must be rationally driven by the specific pharmacokinetic properties of the targeting molecule to maximize therapeutic outcomes. As this result is demonstrated specifically with rhPSMA-10.1, the absolute values presented for therapeutic index and tumor or normal tissue absorbed dose will not be exactly replicated with other radiopharmaceuticals. However, the resulting comparisons between ^225^Ac and ^212^Pb are likely generalizable to any radiopharmaceutical with long retention of the vector in tumors relative to normal tissue.

## Data Availability

All data produced in the present study are available upon reasonable request to the authors

## STATEMENTS & DECLARATIONS

Benjamin Fongenie, Nathaniel Scott and Daniel Stevens are employees of and have stock options/shares in Blue Earth Therapeutics, a clinical stage radiopharmaceutical development company developing beta and alpha labelled radiopharmaceuticals.

AMDB was supported by the National Measurement System of the UK Government’s Department for Science, Innovation and Technology, and by the project 22HLT03 AlphaMet, funded from the European Partnership on Metrology, co-financed from the European Union’s Horizon Europe Research and Innovation Programme and by the Participating States, where the National Physical Laboratory is funded by UKRI under the UK Government’s Horizon Europe Guarantee scheme (no. 10117403).

No other conflicts of interest exist.

All authors contributed to this modelling through review and second checking of raw data and dosimetry calculations. Benjamin Fongenie performed the dosimetry calculations in the first instance. Daniel Stevens and Nathaniel Scott contributed both to the generation of the original ^177^Lu dosimetry data and review of the manuscript.

The ongoing phase I/II study ^177^Lu-rhPSMA-10.1 (NCT05413850) has been performed in line with the principles of the Declaration of Helsinki and was approved by appropriate ethics committees in the US and Netherlands, including Advarra and East Netherlands MEC.

## ACKNOWLEDGMENTS

We acknowledge the hard work of the LabPlot team for enabling data analysis and curve fitting:

LabPlot Team (2025), LabPlot: A FREE, open source, cross-platform Data Visualization and Analysis software accessible to everyone and trusted by professionals, (Version 2.11.1) [Computer software]. https://labplot.org

## KEY POINTS

### QUESTION

Which alpha-emitter should a novel PSMA targeting RLT utilize to maximize absorbed dose delivery to tumors relative to normal tissues such as the kidney?

### PERTINENT FINDINGS

Simple adjustments to effective half-lives in tissue from dosimetry data acquired from a novel PSMA targeting radioligand radiolabeled with an easily imaged radionuclide (^177^Lu) allows theoretical time-activity curves to be plotted for the scenario in which it is labelled with an alpha-emitter. Dosimetry calculations using these time-activity curves and existing data on daughter translocation from ^225^Ac and ^212^Pb indicate ^225^Ac offers superior absorbed dose distribution than ^212^Pb.

### IMPLICATIONS FOR PATIENT CARE

Utilizing ^255^Ac as an alpha-emitter is likely to deliver an improved balance of absorbed dose to tumors relative to normal healthy tissue than ^212^Pb. This may improve clinical outcomes when developing rhPSMA-10.1.

## REFERENCES

1. Kratochwil C, Bruchertseifer F, Giesel FL, et al. 225Ac-PSMA-617 for PSMA-targeted α-radiation therapy of metastatic castration-resistant prostate cancer. Journal of Nucl. Med., 2016:57, 1941–1944,.

2. Yeong CH, Cheng MH, Ng KH., J Zhejiang. Therapeutic radionuclides in nuclear medicine: current and future prospects. Univ Sci B., 2014:15.

3. Abou-Jaoudé, W., & Dale, R. A theoretical radiobiological assessment of the influence of radionuclide half-life on tumor response in targeted radiotherapy when a constant kidney toxicity is maintained. Cancer Biotherapy and Radiopharmaceuticals, 2004:1.

4. C. Stokke, M. Kvassheim, J. Blakkisrud Radionuclides for Targeted Therapy: Physical Properties. Molecules, 2022:27

5. Su F.M., Beaumier P., Axworthy D., Atcher R., Fritzberg A. Pretargeted radioimmunotherapy in tumored mice using an in vivo 212Pb/212Bi generator. Nucl. Med. Biol., 2005.

6. Matthew R. Griffiths, David A. Pattison, Simon G. Puttick et al. First-in-Human 212Pb-PSMA–Targeted α-Therapy SPECT/CT Imaging in a Patient with Metastatic Castration-Resistant Prostate Cancer. Journal of Nuclear Medicine, 2024

7. James Nagarajah, Hyun Kim, Joseph Osborne, et al. Organ and tumour dosimetry of 177Lu-rhPSMA-10.1, a novel PSMA-targeted therapy: results from a Phase I trial. EJNMMI, 2025.

8. Adam L Kesner, Lukas M Carter, Wesley E Bolch et al. MIRD Pamphlet No. 28, Part 1: MIRDcalc—A Software Tool for Medical Internal Radiation Dosimetry. Journal of Nucl. Med., 2023, Vol. 64.

9. Oleksandra V. Ivashchenko, Gerhard Glatting et al. Time-Activity data fitting in molecular Radiotherapy: Methodology. Physica Medica, 2023.

10. M. Li, N. J Baumhover, M. Schultz et al. Preclinical Evaluation of a Lead Specific Chelator (PSC) Conjugated to Radiopeptides for 203Pb and 212Pb-Based Theranostics. Pharmaceutics, 2023:15(2).

11. S. Mirzadeh, K. Kumar, O. Gansow The chemical fate of 212Bi-DOTA formed by β-decay of 212Pb(DOTA)2-, Radiochimica Acta, 1993:60

12. M. Kvassheim, E. Hernes, C. Stokke et al. Clinical phase 0 trial of 212Pb-PSMA therapy AB001: activity concentrations of 212Pb, Athens : European Federation of Medical Physicis, 2023.

13. Zitzmann-Kolbe, S. Biodistribution and contribution of in vivo generated daughter radionuclides to the antitumour afficacy of 225Ac-PSMA-Trillium. Hamburg : EANM, 2024.

14. EANM ‘24 Abstract Book Congress Oct19-21, 2024, DOI: 10.1007/s00259-024-06838-z

15. M. Kvassheim, C. Stokke The Impact of Assumptions on Kidney Absorbed Doses When Modelling Redistribution of Daughters for Alpha-Emitter Based Somatostatin Receptor Therapies. Hamburg: EANM 2024.

16. Z. Ells, T. Grogan, J. Calais et al. Dosimetry of [^177^Lu]Lu-PSMA-Targeted Radiopharmaceutical Therapies in Patients with Prostate Cancer: A Comparative Systematic Review and Metaanalysis. J Nucl Med., 2024:65

17. G. Liubchenko, G. Boning, A. Delker et al Image-based dosimetry for [^225^Ac]Ac-PSMA-I&T therapy and the effect of daughter-specific pharmacokinetics. EJNMMI, 2024:51

18. J. Dos Santos, M. Schafer, C. Kratochwil et al Development and dosimetry of Pb/ Pb-labelled PSMA ligands: bringing “the lead” into PSMA-targeted alpha therapy? EJNMMI 2019:46

19. Strosberg JR, Naqvi S, Maluccio MA et al. Long-term follow-up of peptide receptor radionuclide therapy (PRRT)-naïve patients with gastroenteropancreatic neuroendocrine tumors (GEP-NETs) treated with targeted alpha therapy 212Pb-DOTAMTATE in the Phase 2 ALPHAMEDIX 02 trial. Presented at: European Society for Medical Oncology (ESMO); 2025 Oct 20; Lugano, Switzerland

20. ICRP Report 60, ICRP 1990

21. Barendsen, G.W. The Relationships between RBE and LET for Different Types of Lethal Damage in Mammalian Cells: Biophysical and Molecular Mechanisms. Radiation Research Society, 1994:139.

22. Sgouros, George. MIRD Pamphlet No. 22 - Radiobiology and Dosimetry of Alpha-Particle Emitters for Targeted Radionuclide Therapy.J Nucl Med., 2010, Vol. 15.

23. Robert F Hobbs, Roger W Howell, George Sgouros et al. Redefining Relative Biological Effectiveness in the Context of the EQDX Formalism: Implications for Alpha-Particle Emitter Therapy. Radiat. Res., 2014:181.

24. Jean-Pierre Pouget, Julie Costanzo. Revisiting the Radiobiology of Targeted Alpha Therapy.Montpellier : Front. Med., 2021, Vol. 8.

25. Guerra Liberal et. Belfast. Differential responses to 223Ra and Alpha-particles exposure in.: Frontiers in Oncology, 2022.

26. D. Stevens, B. Waldron, C. Foxton et al Preclinical evaluation of 225Ac-rhPSMA-10.1, a novel radiohybrid PSMA compound for targeted alpha therapy of prostate cancer. EANM, Barcelona, 2023

