## Supplemental Data for "Dosimetry-Driven Alpha Emitter Selection for Radioligand Therapy: Is daughter translocation a significant safety concern?"

**Supplemental Data, EJNMMI – “****Dosimetry-Driven Alpha Emitter Selection for Radioligand Therapy: A Real World Application comparing ^225^Ac with ^212^Pb”**

Benjamin Fongenie^1^, Nathaniel Scott^1^, Nicolas Chouin^2,3^, Ana M. Denis-Bacelar^4^, Daniel J Stevens^1^

^1^Blue Earth Therapeutics Ltd., Oxford, UK;

^2^Nantes Université, Univ Angers, INSERM, CRCI2NA,F-44000 Nantes, France;

^3^Oniris, 44300 Nantes, France;

^4^National Physical Laboratory, Hampton Road, Teddington, UK

| Kidney | | | | | | |
| --- | --- | --- | --- | --- | --- | --- |
| **Model Tested** | **Starting Values** | **Iterations** | **Tolerance** | **AIC** | **F-test** | **Visual Fit?** |
| *A*exp-(λ1+λphys) - A*exp-(λ2+λphys)* | A = 0.5, L1 = 0.1, L2=0.2 | 10000 | 0.0001 | -11.7 | 0.253 | Poor |
| *A*exp(-(λ+λphys))* | A = 3.3, L = 0.012 | 10000 | 0.0001 | 2.24 | 89 | Good |
| *A*exp(-(λ+λphys))+A2*exp(-(λ+λphys))* | A=3.3, L1 = 0.01, A2 = 0.0001 | 10000 | 0.0001 | 2.34 | 47.9 | Good |
| Salivary | | | | | | |
| **Model Tested** | **Starting Values** | **Iterations** | **Tolerance** | **AIC** | **F-test** | **Visual Fit?** |
| *A*exp-(λ1+λphys) - A*exp-(λ2+λphys)* | A = 0.4, L1 = 0.012, L2=1.19 | 10000 | 0.0001 | -15.5 | 42.4 | Good |
| *A*exp(-(λ+λphys))* | A = 0.36, L = 0.012 | 10000 | 0.0001 | -16.3 | 96.1 | Good |
| *A*exp(-(λ+λphys))+A2*exp(-(λ+λphys))* | A=0.35, L1 = 0.015, A2 = 0.0001 | 10000 | 0.0001 | -19 | 103 | Good |
| Tumour | | | | | | |
| **Model Tested** | **Starting Values** | **Iterations** | **Tolerance** | **AIC** | **F-test** | **Visual Fit?** |
| *A*exp-(λ1+λphys) - A*exp-(λ2+λphys)* | A = 6.9, L1 = 0.0012, L2=0.4 | 10000 | 0.0001 | 9.75 | 10.8 | Good |
| *A*exp(-(λ+λphys))* | A = 6.9, L = 0.004 | 10000 | 0.0001 | 19.3 | 0.856 | Poor |
| *A*exp(-(λ+λphys))+A2*exp(-(λ+λphys))* | A=5.9, L1 = 0.004, A2 = 0.7 | 10000 | 0.0001 | -19 | 103 | Poor |

Table 1: Statistical results of curve-fitting for Kidney, Salivary and Tumor time-activity curves from ^177^Lu-rhPSMA-10.1

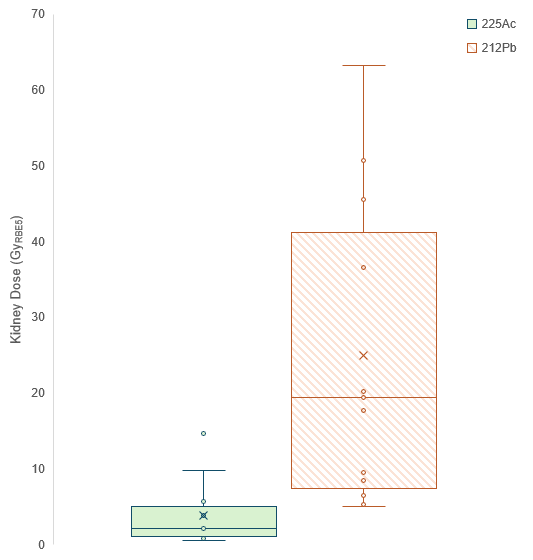

**Fig 1**: Modelled absorbed dose delivered to kidney for 5Gy_(RBE5)_ tumor dose including contributions from translocation of daughters assuming 36% demetallation of ^212^Pb
